# Automated clinical conversations across the cataract pathway with an artificial intelligence (AI) conversation agent: a UK regional service evaluation protocol

**DOI:** 10.1101/2023.06.14.23291399

**Authors:** Automated cataract surgery telephone consult evaluation collaboration, Aisling Higham

## Abstract

**Introduction:** Digital technologies have the potential to support clinical pathways. This study aims to evaluate the impact of using an artificial intelligence-based conversational assistant known as Dora in the cataract pathway. Dora conducts clinical conversations with patients over a telephone call both before and after cataract surgery. Through automation of routine activity, the aim is to increase efficiency of the cataract pathway and the capacity of organisations.

**Method and Analysis:** We will use a mixed-methods cohort-based approach across all sites using Dora in South East England. The study has 3 key objectives, to: 1) Report site-specific variation on the implementation and impact of using Dora 2) assess the impact on the triple bottom line (financial, social and environmental performance) through implementation of Dora 3) understand the real-world patient outcomes of using Dora in clinical pathways. The Dora platform prospectively records symptom and outcome information from each call. We will retrospectively collect data from the hospital record and also collect qualitative data regarding the ease of implementation and patient acceptability of the technology.

**Ethics and dissemination:** This will be registered as a service evaluation at each of the participating clinical sites. Research ethics is not needed as per Health Research Authority guidelines. Site-specific reports will be provided to each participant site as well as an overall report to be disseminated through NHS-England. Results will be published in a formal project report endorsed by stakeholders, and in peer-reviewed scientific reports.

**Article Summary:** *Strengths and limitations of this study:* Strengths

- This is the largest study on the use of an AI based natural language clinical assistant across multiple different hospital sites, with varied geographical locations, demographics and baseline clinical pathways.
- This is a large-scale evaluation with input from multiple independent clinicians, patients, evaluators, economists and strategists.
- Standardised data collection from autonomous clinical assistant Limitations

- Retrospective collection of hospital level follow up data

## Introduction

Healthcare systems in the UK and globally are facing a workforce crisis (The King’s Fund 2022). An increasing number of artificial intelligence (AI) based solutions from image analysis algorithms to conversational agents (Milne-Ives et al. 2020) have the potential to support healthcare staff by freeing clinicians to focus on more complex work, whilst increasing accessibility to healthcare services (Benjamens, Dhunnoo & Meskó 2020).

Dora (Ufonia Ltd, Oxford, UK) is an AI-enabled UKCA-marked autonomous clinical assistant that is capable of conducting natural language phone conversations with a patient throughout the cataract surgery perioperative period. The solution is designed to be accessible with the conversations being delivered through a phone call to any number. For patients, the experience can be comparable to speaking to a doctor or a nurse on the phone, and no apps, devices, or even a mobile phone is needed. Previous studies have been conducted for Dora’s safety and acceptability following routine cataract surgery follow up (de Pennington et al. 2021). Dora calls the patient and has a conversation at 3-4 weeks following routine cataract surgery, and patients with concerns or symptoms receive a call back by a clinician in 48 hours (Khavandi et al. 2022). Previous research evidence shows good levels of patient acceptability, safety and the potential to reduce routine clinical workload by up to 60% (Khavandi et al. 2022).

Ophthalmology is the busiest outpatient specialty in the NHS - it provides over 7.5 million outpatient appointments a year, and cataract surgery is the most common operation worldwide. NHS-England (NHSE) have set targets to reduce outpatient follow up by 25% (NHS England and NHS Improvement 2022) and the promise of automation of a ‘high-volume, low complexity’ (HVLC) pathway such as cataract surgery with AI represents an opportunity to free up workforce and outpatient clinic space. The South East of England (SE) region has identified the cataract pathway as a key area for improvement, with long waiting lists exacerbated by Covid-19, and has commissioned the use of Dora in up to 6 hospital sites for a 12-month evaluation. This approach supports the NHS Long Term Plan goals of increasing “out-of-hospital” and “digitally enabled” outpatient care (NHS 2019).

The aim of this service evaluation is to assess the impact of automating consultations previously conducted by healthcare professionals in the cataract pathway across varied hospital sites and clinical pathways in the SE region. We will use a prospective, mixed-methods cohort-based approach.

The objectives of this study are to describe:

1. Variations in existing clinical pathways for cataract follow up prior to the implementation of Dora;
2. Real-world patient acceptability of Dora across the SE region;
3. Real-world impact on operational efficiency from implementation and variations across sites;
4. Patient outcomes (percentages of patients seen face to face, rates of unplanned care episodes);
5. Variations in access to follow up care based on location, demographic, and clinical traits;
6. Impacts of implementation on the triple-bottom line (social, environmental, financial) (Johnson et al. 2021);
7. Ease of implementation;

The outcomes of this evaluation will inform further development of autonomous AI implementation across the care pathway regionally and nationally.

## Methods and Analysis

### Sample selection

A proportion of Dora calls in BOB (Berkshire, Oxfordshire and Buckinghamshire) and Frimley Integration Care Systems (ICS) are funded via a grant from the Small Business Research Initiative (SBRI). NHSE has also commissioned the use of Dora calls in South East (SE) England. All SE hospitals providing cataract follow-up have been offered the opportunity to adopt this solution. Recruitment was offered via electronic mailing, ICS level presentations, NHS contacts and professional connections. Sites were chosen by the NHSE team and were selected to be both geographically representative of the region, and to be maximally able to benefit from the technology (i.e. focused on those sites with high patient volumes or high waiting lists that could benefit from automation). All sites having Dora calls will be included in the evaluation. It is expected that all clinical sites will have started live patient calls by July 2023.

### Interventions to be measured

Dora R3 (Ufonia Limited, Oxford, UK) is a UKCA-marked clinical conversational assistant that utilises speech recognition and natural language processing to have natural voice conversations with patients over the phone (Gardiner et al. 2022).

Dora will call patients at several time points across the cataract pathway in this evaluation. At each participating site, Dora will have conversations with patients for at least one of the key timepoints (Table 1 and Figure 1).

**Table 1.** The three conversations delivered by Dora.

|  | **Pre-assessment** | **Pre-surgery reminder** | **Post-operative** | **PROMS** |
| --- | --- | --- | --- | --- |
| Timing | After referral to the cataract clinic, but before being listed for surgery | Within the week before planned surgery | 3-4 weeks after routine, uncomplicated cataract surgery | 8-12 weeks after surgery |
| Key outputs | Cataract symptoms and desire to proceed with surgery<br>Medical risk factors<br>Likely ability to tolerate local anaesthetic surgery | Confirm patient's intention to attend surgery date<br>Responses to 5 questions in CatProm5 (Sparrow et al. 2018) | Any significant symptoms (red eye, pain, visual concerns, floaters, flashing lights)<br>Any unanswered clinical questions<br>Recommendation regarding next steps (second eye surgery, further review, discharge) | Responses to 5 questions in CatProm5 (Sparrow et al. 2018) |

PROMS: Patient reported outcome measures

**Figure 1:**
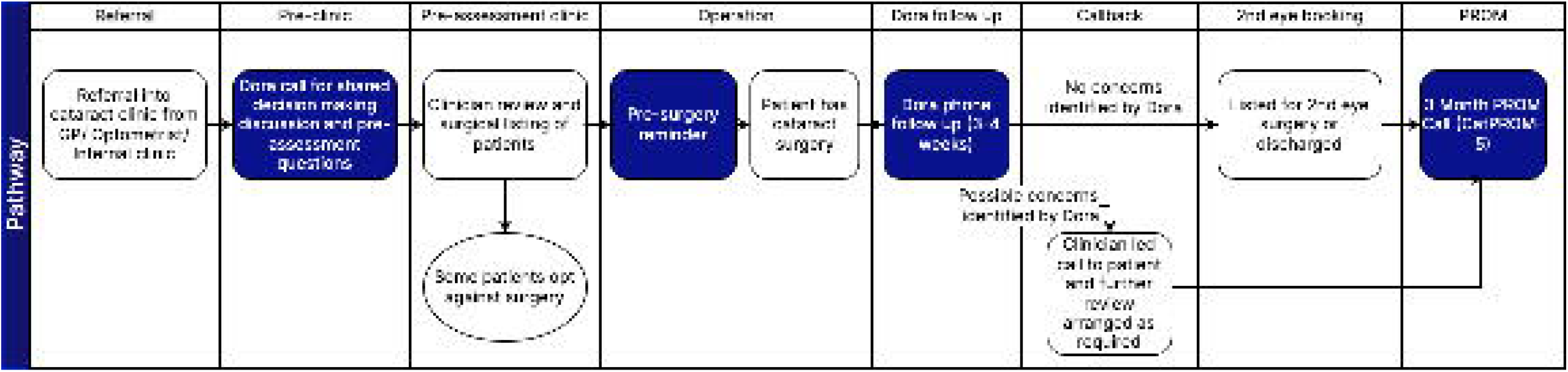
The Standard Cataract pathway with Dora calls.

- **Upon receiving cataract referral -** prior to cataract pre-assessment clinic
- **Prior to surgery**- to collect pre-operative patient reported outcome measures (PROMS) and to confirm the patient plans to attend surgery
- **Post-operatively -** 3 to 4 weeks after cataract surgery, to assess symptoms, address questions and confirm whether 2nd eye surgery is required
- **8 to 12 weeks after cataract surgery**- to collect structured post operative PROM information

Hospital sites add patients suitable for Dora calls onto a clinic list via their existing patient administration system (PAS). This list, with associated demographic data to initiate a call, is received by the Dora platform either via direct integration, or by secure file transfer. Patients can speak to Dora on any landline or mobile phone number recorded in the hospital patient administration system.

For all conversations, Dora’s intended use is for adult patients who would be suitable for a telephone-based call with an equivalent health professional conducted in English. For the pre-operative calls, any patient referred for consideration of cataract surgery is eligible for a call. For the post operative calls, eligible patients are those with routine, uncomplicated cataract surgery who are able to have a telephone call with a health professional. From the patient’s they receive the Dora call on either a mobile or landline number, and speak to Dora like they would to a clinician; no training is required.

The outputs from the Dora call are sent in a spreadsheet via secure file transfer from Ufonia to the hospital for further review. The spreadsheet contains a summary of the patient’s responses to the key symptom or outcome based questions as well as an overall recommendation for onward care (e.g. recommend further clinician review or discharge). A clinician-led call-back is arranged if Dora identifies potential concerns. Table 1 provides an overview of the key outputs of each of the four calls.

The study will last 24 months: a 12 month period during which each site will go live (staggered) and a 3 month period of post evaluation analysis.

### Anticipated Recruitment

According to the National Ophthalmology Database (NOD) report 2022, most centres perform between 1000-3000 cataract surgeries annually (Donachie & Buchan 2022). Previous implementation of Dora has shown that approximately 80% patients meet the eligibility criteria for a Dora call as part of their standard care pathway. We anticipate at least 7 different centres to use Dora in the care pathway, thus including approximately 8400 patients over the 12 month period of this evaluation. Each patient would potentially have four conversations with Dora (pre-assessment, pre-surgery reminder, post-operative, PROMS).

### Patient and Public Involvement

A patient representative who has been involved in the design and conduct of this evaluation and sits on the steering committee. Wider groups of patients have also been instrumental in the design of the Dora product with feedback from testing and focus groups incorporated into the platform (Khavandi et al. 2022). The evaluation has been designed to include assessment of key concerns raised by patient groups previously, particularly regarding the safety of implementing Dora in a real-world setting.

### Data Statement

Technical appendix, statistical code, and dataset will be available from the Dryad repository after collection.

### What outcomes will be measured, when and how

Data will be captured from the clinical sites as they roll out using Dora. This will be compared to data collected before Dora’s implementation. Not every work package will run from every site. Full details are shown in Table 2.

**Table 2.**
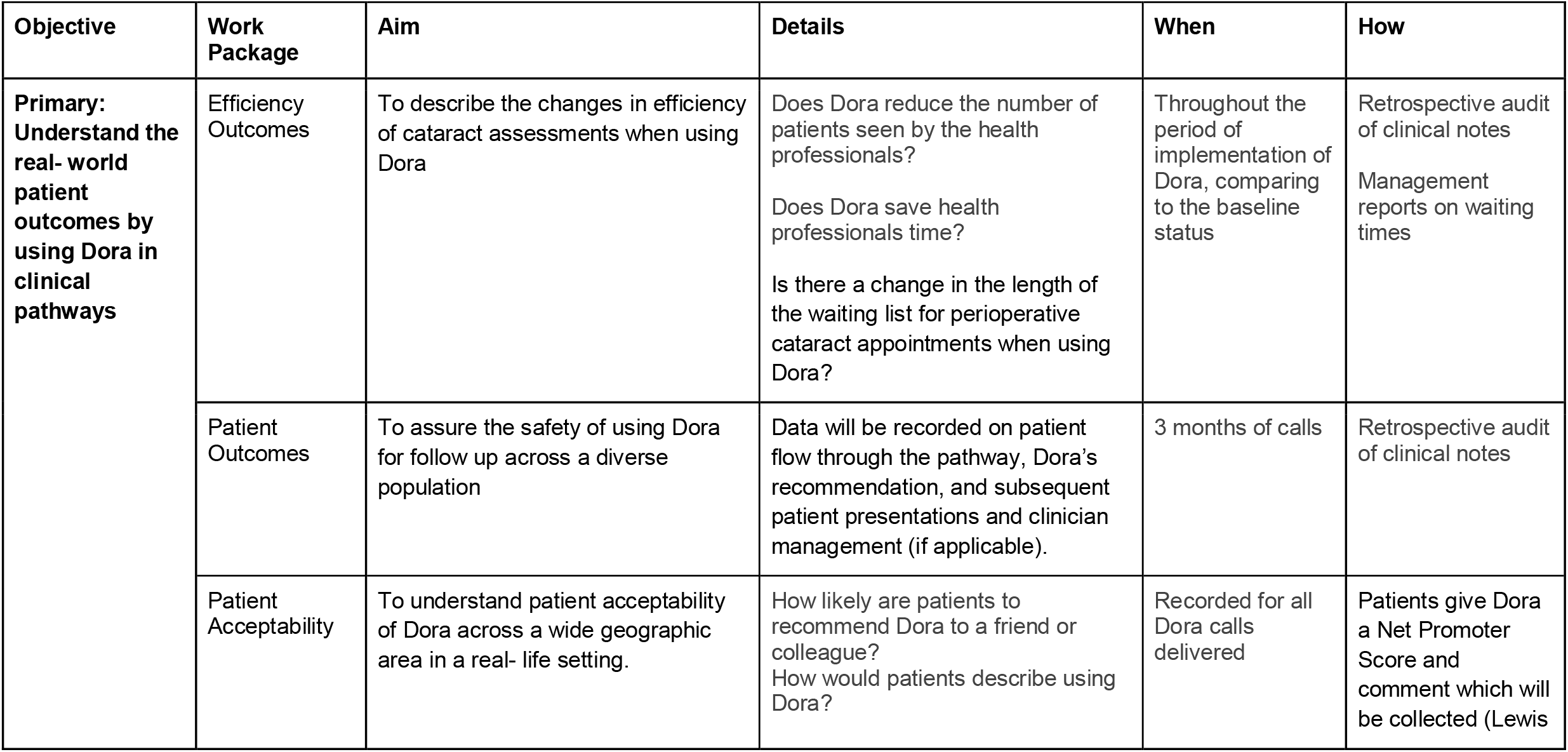
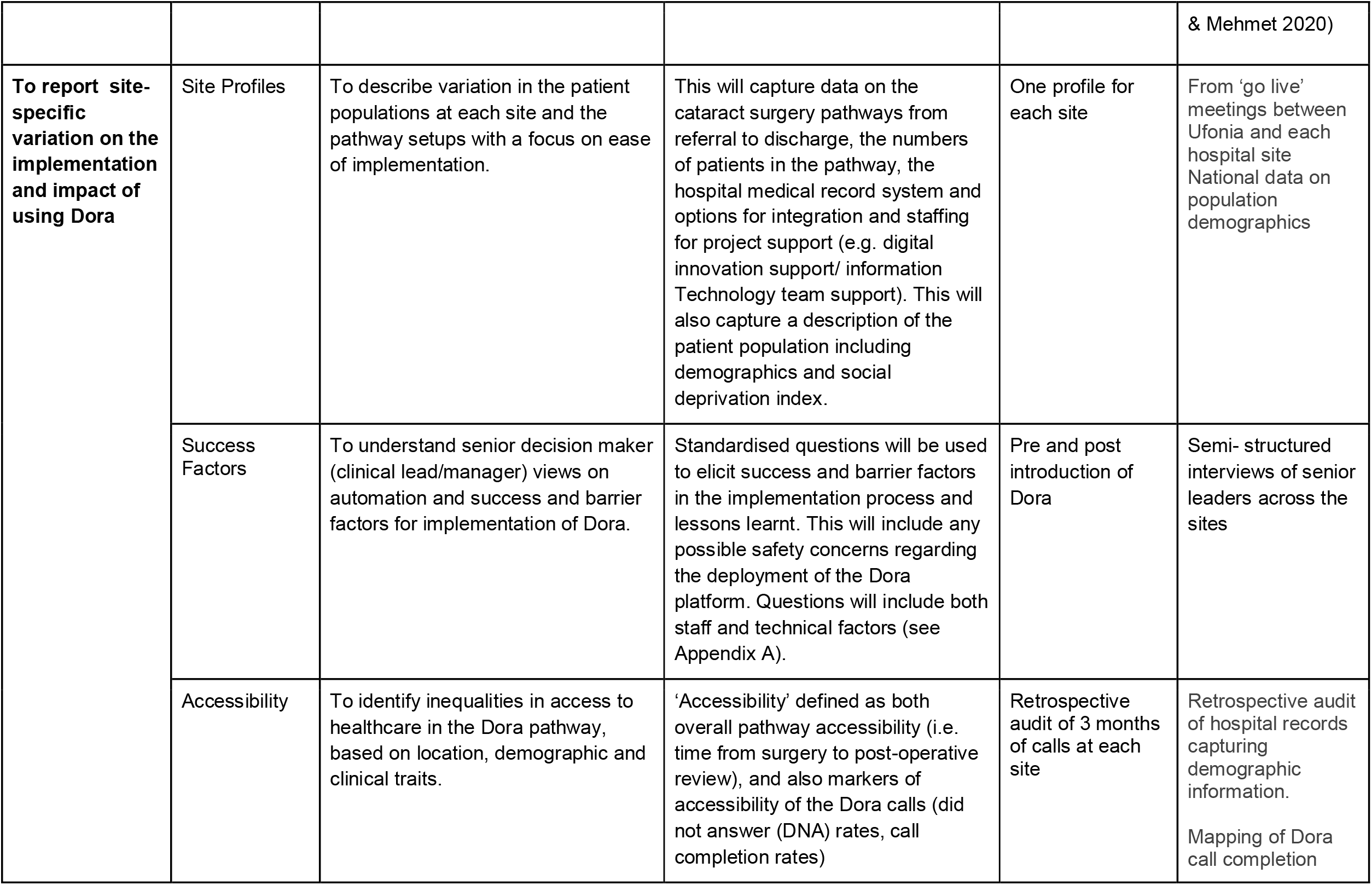
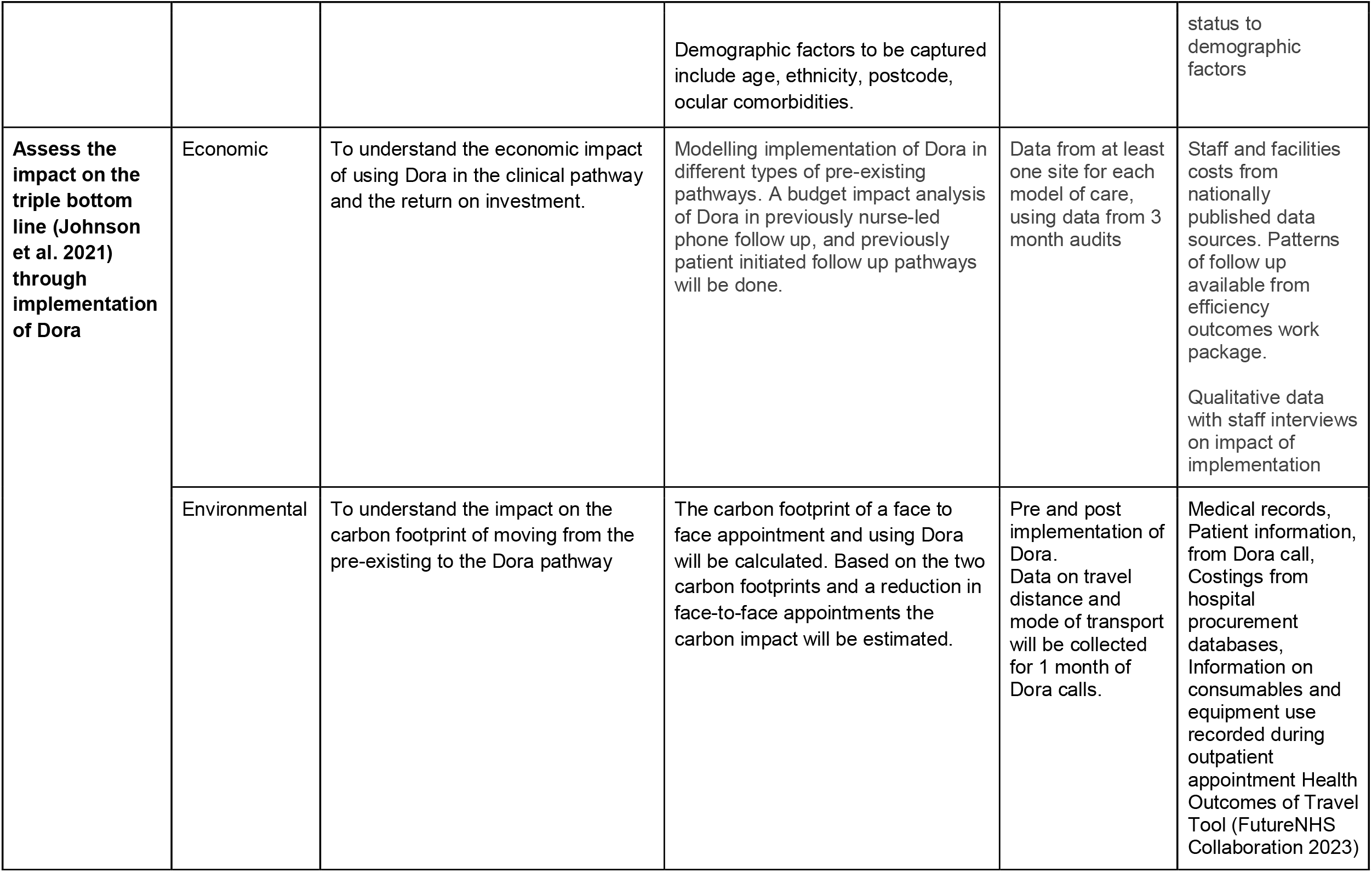
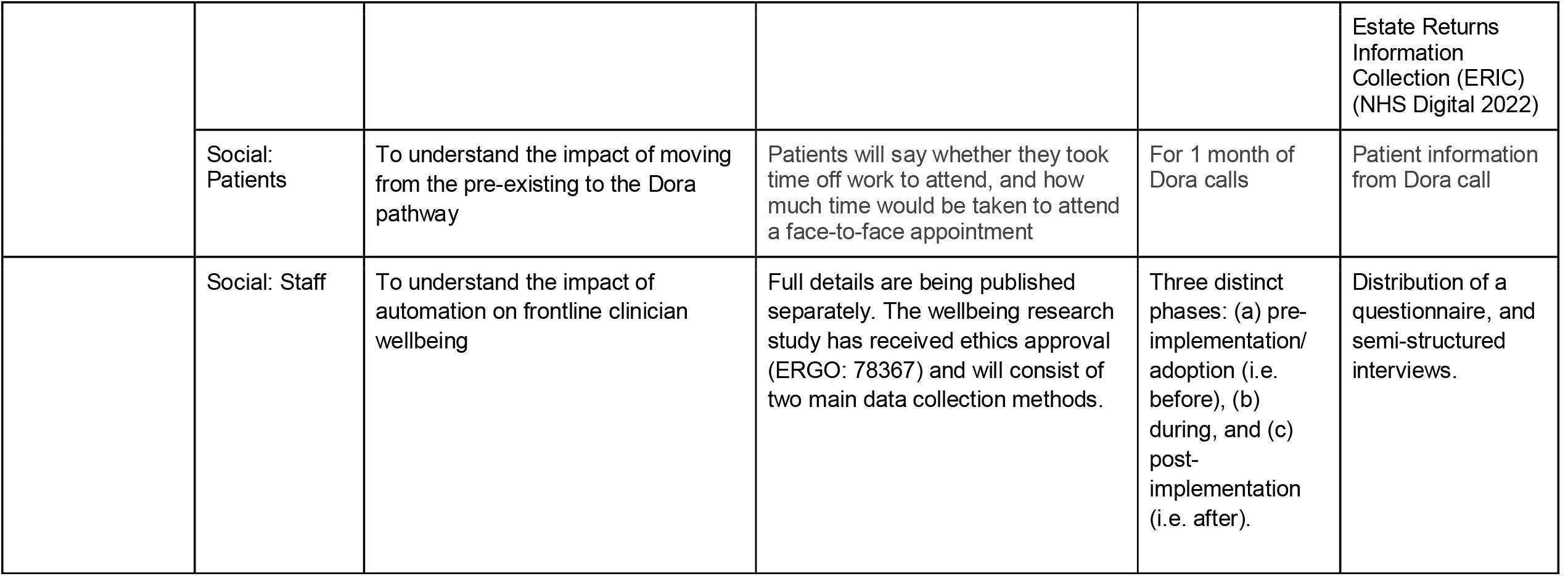
shows the key objectives for this study.

### Data analysis plan

This evaluation will collect data from multiple contributors. The full details of how data of each work package will be analysed is shown in Table 3.

**Table 3:** Details of data analysis plan for each work package.

| **Work Package** | **Data analysis plan** |
| --- | --- |
| **Efficiency outcomes** | Descriptive statistics of patient outcomes after the Dora call including call completion rates.<br>This data will be subgrouped by age, ethnicity and location. |
| **Patient Outcomes** | Descriptive statistics on patient flow through the pathway. This will include Dora's recommendation regarding next steps in care, and subsequent patient presentations and clinician management (if applicable). |
| **Patient Acceptability** | Calculated Net Promoter Score (NPS) (Lewis & Mehmet 2020)<br>Thematic analysis of NPS comments from patients |
| **Site Profiles** | Descriptive report on site characteristics |
| **Success factors** | Thematic analysis of interviews to draw out the key themes. |
| **Accessibility** | Descriptive report on patients using Dora including key demographics (age, gender, ethnicity). Comparison of whether there is a significant difference between patient Dora call outcomes based on demographic factors. |
| **Economic** | Economic model of the whole cataract pathway. Using data from the efficiency work package, and applying logistic regression analysis with respect to Dora call outcomes taking various explanatory factors (such as demographics) into account. |
| **Environmental** | To estimate the carbon footprint of a face to face outpatient appointment and an automated phone triage a hybrid carbon footprinting method will be used. Where possible the carbon footprint will be based on a process-based carbon footprint analysis, complemented by environmentally extended input-output analysis where data is not available. The model will detail carbon footprint locally, regionally and nationally. |
| **Social** | Patients: Descriptive statistics of patient time required to attend a face-to- face outpatient clinic.<br>Staff: The semi structured interviews will be recorded, transcribed and analysed using thematic analysis. The data from the validated surveys will be collected both pre and post implementation and analysed with descriptive statistics. |

### Statistical analysis

In relation to the Accessibility theme, descriptive statistics of the sociodemographic and clinical profile of each site and the overall patient cohort will be done using proportion for categorical data, mean with standard deviation for parametrically distributed continuous data and median with interquartile range for non-parametric data. This data will also be stratified by demographic data (age, gender, ethnicity, location) and clinical data (ocular comorbidity). Results will be compared using the univariable χ2 test when comparing proportions, student’s t-test for parametric data and kruskal-wallis test for non-parametric data.

In relation to the Economic theme, for comparison between outcomes pre and post the implementation of Dora, we will undertake a logistic regression analysis on the probability of requiring outpatient treatment, taking into account demographic data as explanatory factors.

A two-tailed significance threshold of 0.05 will be used throughout the analysis.

## Trial Registration

This evaluation will be submitted to ClinicalTrials.gov for registration.

## Ethics and confidentiality

All sites will be using Dora as part of their provision of care to patients. To deliver this service, Ufonia is a data processor for Patient Identifiable Data (PID) which is in line with the governance arrangements for all other clinical software applications, and this arrangement forms part of an information processing agreement with the Trust. Per the (Health Research Authority 2023) this study meets all the criteria for both a hospital service evaluation, and a Post Market Surveillance (PMS) service evaluation of a UKCA marked device. Buckinghamshire Healthcare NHS Trust Project Classification Group have reviewed the evaluation and classified it as “Service Review/ Quality Improvement” Reference: PCG138. The clinician wellbeing study has received research ethics approval from the University of Southampton Ethics and Research Governance Online (ERGO: 78367).

All sites must secure local audit approval prior to collecting data. When undertaking the service evaluation, data will be transferred from NHS sites to the data team for analysis via password protected, NHSmail-linked access-controlled files. Prior to transfer and analysis, all data will be de-identified, meaning that no consent will be required from patients for this purpose. To avoid any potential breach of confidentiality, only staff of the participating organisations directly involved in collection of data will have access to health records. No specific recruitment will be performed for these data as only anonymised data collected as part of routine patient care will be used. An independent steering board will meet every six weeks to review progress of the project relating to the key work packages.

### Dissemination

This will be conducted as a regional audit of practice in conjunction with trainee research collaboratives, with support from patient representatives, ophthalmologists, and the NHSE Business Intelligence team.

Site-specific reports will be provided to each participant site as well as an overall report to be disseminated through NHSE. Results will be published in a formal project report endorsed by stakeholders, and in peer-reviewed scientific reports.

## Data Availability

All data produced in the present study are available upon reasonable request to the authors

## Author contributions

Authors’ Contributions: AH, EL and SK conceived the study topic and designed and drafted the proposal. All other authors contributed to drafting and revising the proposal. Final revision was conducted by AH.

## Acknowledgements

We would like to thank the staff at all clinical sites for their ongoing support in implementing Dora calls within the clinical pathways.

## Appendix A

**Questions within the interview prior to the introduction of ‘Dora’**

1. Can you tell me a little bit about your clinical/managerial role?
2. How did you find out about the Dora project?
3. What were your first thoughts when you heard about Dora?
4. What difference are you hoping Dora will make to your service/team/patients as a whole?
  ⍰ Draw out how/if Dora will/will not change
  ⍰ Think about change to pathway change
5. Do you have any thoughts on how staff will respond to the change?
6. Do you have any thoughts on how the patients will respond to the change?
7. What are your key thoughts around the introduction of Dora into your service?
  ⍰ Draw out positive and any concerns
  ⍰ What might affect/have an effect implementation (? Service constraints etc. barriers/enablers)
8. What initial thoughts have you around pathway redesign and who should be involved in the project design
9. Could you let me know of any current opportunities/ challenges in your department?
10. Can you tell me about how you think Dora will support current opportunities, be part of the solution to current challenges?
11. Is this project being funded? If so, how?
  ⍰ Draw out any thoughts about funding going forward post the project
12. How were you approached to be involved in the project? (asked or volunteered).

**Questions within the interview post the introduction of “Dora”**

*These questions are subject to change and will be informed from findings in initial interviews*

1. What has been your experience about Dora being introduced in the service (how embedded)
2. What has made it easier? *Assumptions on what might make it easier*
  ⍰ Information Governance/data sharing (organisational factors)
  ⍰ Staff on board
3. What has made it harder? *Assumptions on what might make it harder*
  ⍰ Information Governance/data sharing (organisational factors)
  ⍰ Staff resistant

**Clinical/managerial lead involvement**

1. What level of involvement was required of you to roll out the project?
2. How much time did you think it would involve compared to actual? (time commitment
  - what has it taken up, what has most of your time been spent on)
3. Did you have any project support locally from your Trust?

**Staff/People factors**

1. How did you introduce the concept to staff?
2. How did you communicate the change to staff?
3. How did you co-designed pathway re-design with staff?
4. Did you trial and test the pathway before the introduction to the service?
5. What training/preparation was there for staff? (timely?)
6. Were any concerns/ideas raised by staff? (how were these addressed)
7. How did you introduce this to patients?
8. How did you decide which patients were eligible?

**Financial factors**

1. Scale-ability – feasibility for continuing at site (post NHSE 1 year funding)

**Lessons learned to overcome barriers (enablers) – use if not enough from Q1, 2 and 3**

1. What have you learned along the way?
2. If you could share one piece of key information for another site looking to implement, what would it be?

